# Ten-year Cost Trends of Rate and Rhythm Control Medications in the United States, 2013-2022

**DOI:** 10.64898/2025.11.28.25341239

**Authors:** Nabeel Sami, Anas Mahmood, Aparna Dintakurti, Michael Fragner

## Abstract

**Background:** Rate and rhythm control medications remain central to atrial fibrillation (AF) management, even as catheter ablation becomes increasingly preferred for select patients. However, national spending patterns and cost disparities between these medication classes under Medicare Part D remain poorly defined.

**Objectives:** To evaluate Medicare Part D spending trends and cost differences between rate and rhythm control agents from 2013 to 2022.

**Methods:** A retrospective analysis of Medicare Part D Prescription Drug Event data (2013–2022) was performed, including 13 commonly prescribed monotherapy agents categorized as rate or rhythm control. For each drug, total annual spending, cost per claim, and cost per beneficiary were calculated. Linear regression assessed temporal cost trends, and independent *t*-tests compared rate versus rhythm classes. Statistical significance was defined as *p* < 0.05.

**Results:** Rate control agents showed a significant decline in cost per claim (*p* = 0.003), driven by metoprolol succinate (−53.9%, *p* < 0.001). Rhythm control agents demonstrated significant increases in both cost per claim (*p* < 0.001) and cost per beneficiary (*p* = 0.026). Dronedarone rose from $2,166 to $5,374 per beneficiary (*p* < 0.001), while flecainide increased to $71 per claim and $276 per beneficiary (*p* = 0.001, *p* = 0.036). Despite generic availability, amiodarone, verapamil, and diltiazem also trended upward. Across all years, rhythm agents remained more expensive than rate agents (*p* < 0.001).

**Conclusion:** From 2013–2022, Medicare costs decreased for rate control medications but rose sharply for rhythm agents. Addressing these disparities through transparent pricing and formulary reform is essential to preserve equitable access to rhythm control therapy.

## Introduction

Atrial fibrillation (AF) is the most common sustained arrhythmia and a leading contributor to cardiovascular-related healthcare spending, particularly among older adults. Management typically follows one of two pharmacologic strategies, rate control or rhythm control, both endorsed by contemporary guidelines. Although these therapeutic pathways are well established, their long-term economic implications for patients, especially those covered under Medicare, remain insufficiently characterized.

Over the past decade, catheter ablation has become increasingly favored as a first-line rhythm control therapy for symptomatic AF, particularly in patients with paroxysmal disease. Nevertheless, a substantial proportion of patients continue to rely on pharmacologic therapy due to procedural risk, comorbidities, or limited access to specialized centers. Consequently, rate- and rhythm-controlling medications remain central to real-world AF management, particularly among Medicare beneficiaries.

Recent prescribing trends show growing utilization of both legacy generic agents and newer, high-cost antiarrhythmics. Commonly used generics such as metoprolol and amiodarone remain staples of therapy, while higher-cost agents like dronedarone and flecainide have gained traction as alternatives, often with minimal cost transparency for patients and payers. These evolving utilization patterns raise important questions about the trajectory of national spending and cost disparities between drug classes, especially as early rhythm control gains greater clinical emphasis.

This study evaluates national Medicare Part D trends in cost per claim and cost per beneficiary from 2013 to 2022 for 13 widely prescribed rate and rhythm control medications used in atrial fibrillation management. By quantifying cost trajectories across therapeutic classes, this analysis aims to inform healthcare policy and optimize access to evidence-based AF therapies.

## Methods

We conducted a retrospective observational analysis using publicly available Medicare Part D Prescription Drug Event data from 2013 to 2022, encompassing over 2.5 million beneficiaries per year who received at least one rate or rhythm control medication. Medications were selected based on clinical relevance to atrial fibrillation management, focusing on agents routinely prescribed as part of rate or rhythm control strategies. Thirteen monotherapy agents were included and categorized into rate control (metoprolol tartrate, metoprolol succinate, atenolol, carvedilol, diltiazem, verapamil, and digoxin) and rhythm control (flecainide, propafenone, dofetilide, amiodarone, sotalol, and dronedarone) classes. Combination therapies and non-monotherapy agents were excluded.

For each medication, three annual spending metrics were derived: (1) total annual Medicare Part D spending, (2) cost per claim (total annual spending divided by number of claims), and (3) cost per beneficiary (total spending divided by the number of unique patients receiving the medication). These values were computed for each calendar year and aggregated by therapeutic class. Pooled averages were calculated for rhythm control versus rate control agents to enable class-level comparisons across the study period.

To assess longitudinal cost trajectories, ordinary least squares (OLS) linear regression models were used with year as the independent variable and cost per claim or cost per beneficiary as dependent variables. Regression analyses were conducted for each drug individually to evaluate annual cost trends. Additionally, independent-sample *t*-tests were performed to compare average cost per claim and cost per beneficiary between pooled rhythm and rate control groups across all study years. Statistical significance was defined as a two-tailed *p* < 0.05.

All analyses and visualizations were performed in Python (Google Colab environment) using validated scripts. Outputs were independently reviewed and interpreted by the authors to ensure analytic consistency and accuracy.

## Results

From 2013 to 2022, rate and rhythm control medications demonstrated distinct cost trends. Linear regression showed that rate control agents experienced a statistically significant decline in cost per claim (p = 0.003), while rhythm control agents had a significant increase over time (p < 0.001). When evaluating cost per beneficiary, rhythm control agents showed a significant upward trend (p = 0.026), in contrast to rate control agents that did not show a statistically significant change (p = 0.163). These divergent trends are illustrated in the six-panel plot (Figure 1).

**Figure 1.**
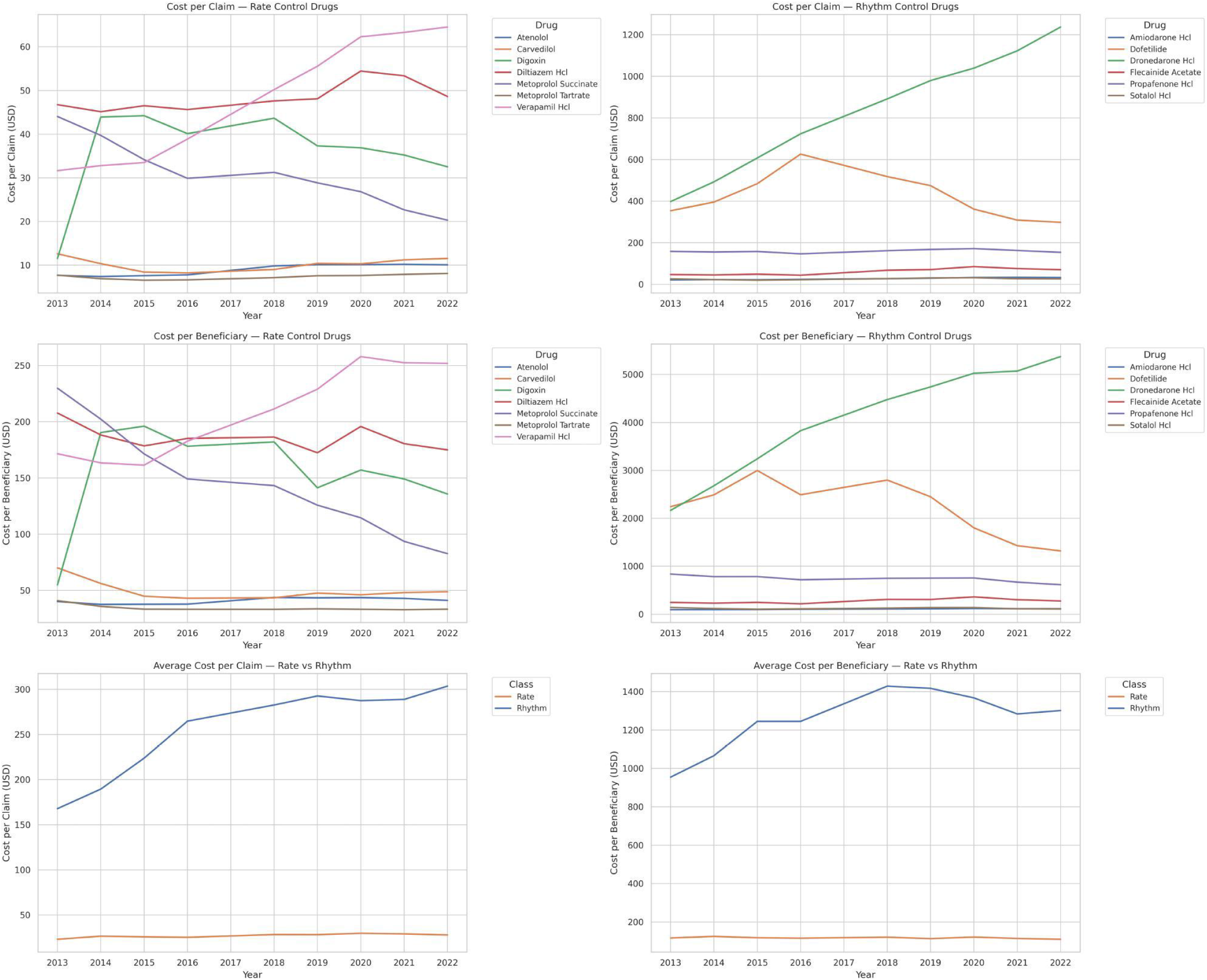
Medicare Part D cost trends for rate and rhythm control medications from 2013 to 2022. The figure displays cost per claim and cost per beneficiary for individual drugs, as well as average class-level costs over time.

When comparing individual medications, metoprolol succinate demonstrated the most substantial decrease in cost per claim (p < 0.001), decreasing over 50% from approximately $44 in 2013 to $20 in 2022. Metoprolol tartrate also declined, but this trend was not statistically significant (p = 0.054). Among other agents with declining costs, dofetilide demonstrated a statistically significant decrease in cost per beneficiary, falling from $2244 in 2013 to $1320 in 2022 (p = 0.029), although its cost per claim did not significantly change (p = 0.293). Propafenone also showed a significant decrease in cost per beneficiary (p = 0.0025). Carvedilol did not exhibit statistically significant trends.

In contrast, several agents exhibited significant upward cost trends. Dronedarone had the highest cost per beneficiary in 2022 at $5,374, increased from $2166 in 2013, and showed significant increases in both cost per claim and cost per beneficiary (p < 0.001 for both). Flecainide, despite being generic, maintained a high cost and demonstrated statistically significant increases in both cost metrics, reaching $71 per claim (p = 0.001) and $276 per beneficiary in 2022 (p = 0.036). Verapamil showed a significant increase in cost over time, reaching $65 per claim and $252 per beneficiary (p < 0.001 for both). Amiodarone demonstrated a consistent and statistically significant increase in cost, with a 2022 cost of $33 per claim and $115 per beneficiary (p < 0.001 for both). Atenolol increased modestly in cost per claim, reaching $10 (p < 0.001), and cost per beneficiary, reaching $41 in 2022, though this increase was not statistically significant (p = 0.063). Digoxin stood out among the rate control agents because it was inexpensive at the start of the study period, but its cost per claim nearly tripled from $12 in 2013 to $33 in 2022, mirroring the cost of amiodarone, though this trend was not statistically significant (p = 0.689). Sotalol showed no significant change in either cost per claim or cost per beneficiary (p = 0.227 and p = 0.287, respectively). These drug-level cost trends are summarized in Table 1.

**Table 1.** Percent change in Medicare Part D spending for atrial fibrillation medications from 2013 to 2022.

| **Medication** | **Percent Change in Cost per Claim (2013–2022)** | **Percent Change in Cost per Beneficiary (2013–2022)** |
| --- | --- | --- |
| Atenolol | +31.4% | +2.3% |
| Carvedilol | -8.5% | -30.4% |
| Digoxin | +182.4% | +147.6% |
| Diltiazem | +4.0% | -15.8% |
| Metoprolol Succinate | -53.9% | -64.0% |
| Metoprolol Tartrate | +5.1% | -18.7% |
| Verapamil | +104.0% | +46.9% |
| Amiodarone | +53.8% | +22.7% |
| Dofetilide | -15.7% | -41.2% |
| Dronedarone | +210.6% | +148.1% |
| Flecainide | +49.8% | +12.1% |
| Propafenone | -2.8% | -26.6% |
| Sotalol | +1.6% | -22.3% |

Independent two-sample t-tests comparing pooled cost metrics across all years revealed that rhythm control medications were significantly more expensive than rate control medications. This was true for both cost per claim (p < 0.001) and cost per beneficiary (p < 0.001).

Cost trajectories for individual medications and class-level averages are shown in Figure 1, which illustrates changes in cost per claim and cost per beneficiary from 2013 to 2022.

Table 1 shows the percent change in cost per claim and cost per beneficiary for each rate and rhythm control medication from 2013 to 2022.

## Discussion

Metoprolol succinate demonstrated the most substantial cost decline among all agents, with its cost per claim falling by more than 50% from 2013 to 2022. This trend likely reflects expanded generic availability due to increased manufacturer participation, which created meaningful pricing competition. In contrast, flecainide, a rhythm control agent with longstanding generic status, exhibited significant increases in both cost per claim and cost per beneficiary. Despite its generic status, only two manufacturers entered the U.S. market between 2013 and 2022, limiting competition and contributing to persistently high costs.^1^□^2^ This contrast illustrates that generic availability alone does not ensure affordability, particularly with inadequate market competition. Other variables that may contribute to drug pricing include updated clinical guidelines or changing insurance policies.

Rate control medications as a group demonstrated a significant decline in cost per claim over the study period, largely driven by reductions in metoprolol succinate pricing. In contrast, rhythm control medications became more expensive over time in both per-claim and per-beneficiary metrics. Dronedarone had the highest cost per beneficiary by 2022, at $5,374, while flecainide and amiodarone also significantly trended upward in price. The pricing disparity between rate and rhythm control classes may reflect differences in prescribing frequency and barriers such as formulary restrictions or prior authorization.^3^

An important contextual consideration is the increasingly critical and evolving role of catheter ablation in treating atrial fibrillation. Current guidelines now support ablation as a first-line therapy for many patients, particularly those with symptomatic paroxysmal atrial fibrillation and preserved left ventricular function.□ Trials such as CASTLE-AF and CABANA have demonstrated that catheter ablation not only improves rhythm control, but also leads to superior clinical outcomes compared to medical therapy alone. In CASTLE-AF, ablation significantly reduced all-cause mortality and heart failure hospitalizations in patients with reduced ejection fraction, while concurrently improving ejection fraction and quality of life. CABANA showed that ablation reduced major cardiovascular events and resulted in lower rates of atrial fibrillation recurrence.□ Together, these trials support a shift toward ablation as the preferred rhythm control strategy when initiated early. However, not all patients are willing to undergo an invasive procedure or are ineligible due to the clinical risks. As a result, pharmacologic therapy, particularly rhythm control agents, remain essential for a substantial proportion of patients who do not receive ablation or require medications as adjunctive therapy post-procedure. This is especially relevant in patients with heart failure, where a large 2024 cohort study showed that early rhythm control using antiarrhythmics was associated with lower mortality and fewer hospitalizations compared to rate control.□ These findings reinforce the clinical importance of rhythm control medications and highlight the need to ensure their affordability and accessibility.

These data underscore the importance of ongoing evaluation of Medicare Part D formularies to ensure equitable access to both rate and rhythm control agents. Given the expanding evidence base favoring early rhythm control, maintaining cost parity between therapeutic classes may prevent unintentional economic disincentives that skew treatment decisions toward rate control. Formulary adjustments that promote competition, such as encouraging generic manufacturer entry and reducing prior authorization barriers, could optimize both clinical outcomes and healthcare spending efficiency.

While earlier studies like AFFIRM did not show a survival benefit of rhythm over rate control, more recent evidence has suggested otherwise.□ The shift toward earlier rhythm control, as supported by the EAST-AFNET 4 trial, has further increased the clinical relevance of these medications by demonstrating that early rhythm intervention can reduce cardiovascular complications compared to rate control alone.□ Therefore, the observed cost disparities in rhythm agents are concerning, specifically if these rising prices influence clinician prescribing behavior or patient access to needed medications.

Furthermore, amiodarone, another cornerstone rhythm control agent, also showed a significant increase in cost per claim over the study period (p < 0.001), reaching approximately $33 in 2022. Although amiodarone is among the most widely used rhythm control medications, it is often assumed to be low-cost due to its age and generic availability. Our findings reveal that even longstanding, widely used medications may not be insulated from inflationary pressures and shifts in policy or market dynamics. This may suggest that the rising cost of amiodarone may further constrain rhythm control strategies for Medicare beneficiaries, especially if used long term over many years.

While rhythm control agents are often reserved for more selective use, the high and rising costs observed in our study suggest that affordability is increasingly becoming a barrier to optimal care. For Medicare patients with limited income, these trends could discourage adherence to guideline-recommended therapy, exacerbate treatment inequities among older adults, and challenge the sustainability of value-based arrhythmia care.

## Limitations

This study utilized Medicare Part D Prescription Drug Event data, which captures aggregate annual spending but does not provide any information on the clinical indications the medications are used for. Consequently, it is not possible to determine whether the medications analyzed were prescribed specifically for atrial fibrillation or for other conditions such as hypertension or heart failure. This limitation is particularly relevant for rate control agents, which are commonly used across a broad spectrum of cardiovascular diagnoses.

Additionally, the dataset does not include information on medication adherence, prescription refill frequency, or treatment duration, all of which are factors that could influence both cost and utilization patterns. While we focused on 13 commonly prescribed monotherapy agents, combination formulations were excluded, which may have underestimated the total pharmaceutical cost burden associated with arrhythmia management.

Lastly, our findings are limited to the Medicare Part D population and may not fully represent cost trends among patients with private insurance or those without drug coverage. Nonetheless, the national scope, large sample size, and long timeframe provides a strong foundation for examining real-world cost trends and highlighting disparities between rate and rhythm control therapies in the United States.

## Conclusion

There is a widening cost disparity between rate and rhythm control medications used in the management of atrial fibrillation and related arrhythmias. Over the past decade, rate control agents, such as metoprolol succinate, have become increasingly affordable due to robust generic competition and widespread clinical use. In contrast, rhythm control agents remain considerably more expensive and have demonstrated significant price increases over time, regardless of generic availability.

These cost trends have important clinical and health policy implications. While catheter ablation is now supported as a first-line therapy for many patients with symptomatic atrial fibrillation, particularly in light of trials such as EAST-AFNET 4 and CABANA, pharmacologic rhythm control remains essential for patients who are ineligible for ablation, prefer medical therapy, or require adjunctive medications post-procedure. The rising cost of rhythm agents may create access barriers for Medicare beneficiaries and limit the adoption of rhythm-based strategies that have been shown to reduce cardiovascular complications.

To ensure equitable and evidence-based arrhythmia care, policy strategies should promote greater transparency in drug pricing and thoughtful Medicare formulary design that encourages competition without imposing restrictive access criteria. Aligning affordability with evolving guideline-based care will be essential to prevent cost from becoming a barrier to optimal rhythm control, a challenge that, if unaddressed, risks widening disparities in arrhythmia management for older adults.

## Data Availability

All data used in this study are publicly available from the Centers for Medicare & Medicaid Services (CMS) Medicare Part D Prescriber Public Use File. The datasets can be accessed and downloaded directly from the CMS data repository at: https://data.cms.gov/provider-summary-by-type-of-service/medicare-part-d-prescribers/medicare-part-d-prescribers-by-provider-and-drug/data/2022

## Glossary

Not applicable.

## Acknowledgements

None.

## Author Contributions

The authors were solely responsible for study conception, data analysis, manuscript drafting, and final approval.

